# Wastewater surveillance overcomes socio-economic limitations of laboratory-based surveillance when monitoring disease transmission: the South African experience during the COVID-19 pandemic

**DOI:** 10.1101/2024.09.20.24314039

**Authors:** Gillian Maree, Fiona Els, Yashena Naidoo, Laven Naidoo, Phemelo Mahamuza, Mokgaetji Macheke, Nkosenhle Ndlovu, Said Rachida, Chinwe Iwu-Jaja, Setshaba Taukobong, Sibonginkosi Maposa, Kathleen O’Reilly, Mukhlid Yousif, Kerrigan McCarthy

## Abstract

Wastewater and environmental surveillance has been promoted as a communicable disease surveillance tool because it overcomes inherent biases in laboratory-based communicable disease surveillance. Yet, little empirical evidence exists to support this notion, and it remains largely an intuitive, though highly plausible hypothesis. Our interdisciplinary uses WES data to show evidence for underreporting of SARS-CoV-2 in the context of measurable and statistically significant associations between economic conditions and SARS-CoV-2 incidence and testing rates. We obtained geolocated, anonymised, laboratory-confirmed SARS-CoV-2 cases, wastewater SARS-CoV-2 viral load data and socio-demographic data for Gauteng Province, South Africa. We spatially located all data to create a single dataset for sewershed catchments served by two large wastewater treatment plants. We conducted epidemiological, persons infected and principal component analysis to explore the relationships between variables. Overall, we demonstrate the co-contributory influences of socio-economic indicators on access to SARS-CoV-2 testing and cumulative incidence, thus reflecting that apparent incidence rates mirror access to testing and socioeconomic considerations rather than true disease epidemiology. These analyses demonstrate how WES provides valuable information to contextualise and interpret laboratory-based epidemiological data. Whilst it is useful to have these associations established for SARS-CoV-2, the implications beyond SARS-CoV-2 are legion for two reasons, namely that biases inherent in clinical surveillance are broadly applicable across pathogens and all pathogens infecting humans will find their way into wastewater albeit in varying quantities. WES should be implemented to strengthen surveillance systems, especially where economic inequalities limit interpretability of conventional surveillance data.

## Introduction

Surveillance is a core component of the International Health Regulations, and central to the Global Health Security Agenda[1]. World Health Assembly member states are obliged to detect, assess, notify and report events, and to assess their capacity to do so using the Core Capacities document[2]. During the COVID-19 pandemic, which led to an estimated 775 million laboratory-confirmed cases to date[3], testing of individual patient clinical material (usually nasopharyngeal swabs by PCR) and indicators based on these data (including testing rate, incidence rate and proportion testing positive) were the major epidemiological tools used to support monitoring of the pandemic and government decision-making.

SARS-CoV-2 is transmitted by droplet and airborne transmission. Factors predisposing to transmission include crowded conditions, proximity and duration of contact, particularly in the absence of mask-wearing[4]. The clustering of these factors in low socioeconomic households and communities has been shown to exacerbate disease transmission and lead to higher disease incidence[5,6]. However, from the earliest days of the pandemic, it was observed that countries and regions with poorer socioeconomic status reported fewer cases of SARS-CoV-2[7,8]. Most importantly, a reason for this was limited access to testing, evidenced at a global scale by the African region reporting the fewest SARS-CoV-2 cases across the globe alongside the least number of SARS-CoV-2 tests per capita[8].

South Africa, a country with a population of over 62 million persons resident in nine provinces and 52 health districts, experienced five reported waves of SARS-CoV-2[9] each caused by different genetic variants. Following initial detection, the first (ancestral strain) and second waves (Beta variant) of the pandemic occurred between March-June 2020, and from November 2020 to February 2021[10]. The third and fourth waves occurred from May to September 2021 (Delta variant)[10] and from November 2021 to January 2022 (Omicron BA.1 variant)[11]. A subsequent resurgence in cases was responsible for a fifth wave dominated by the Omicron lineage BA.4 and BA.5[10]. The impact of the COVID-19 pandemic was devastating. Disease transmission levels across South Africa were high, with seroprevalence data after the third wave of SARS-CoV-2, suggesting that infections were likely 7.8 times higher than the number of laboratory-confirmed cases[12]. Excess deaths greatly exceeded reported deaths, indicating that official statistics underestimated the death rate[13]. In spite of active case finding[14] and the availability of a comprehensive laboratory network, these data suggested extensive under diagnosis and under-reporting of cases.

These observations foregrounded the intrinsic shortcomings of traditional approaches to laboratory-based communicable disease surveillance programmes. Patient factors (such as health care acceptability and accessibility, financial means to procure testing and the presence and severity of symptoms); health system factors (such as clinician propensity to test, test availability, financial support for testing), and laboratory factors (clerical errors, inherent test performance characteristics) mediate laboratory testing and in turn impact the sensitivity, quality and representivity of surveillance data. Traditional case-based surveillance methods thus underestimate disease burden.

Since the COVID-19 pandemic, wastewater and environmental surveillance (WES) has increasingly been implemented as a complementary surveillance modality that has potential to overcome these limitations[15]. WES has proven utility in supporting polio surveillance by providing highly sensitive data on the presence of poliovirus in communities, material for genomic sequencing and by supporting identification of chains of transmission[16]. These data have greatly enabled polio risk assessments and decision-making regarding public health interventions including the need for vaccination campaigns. During the COVID-19 pandemic, multiple advantages of WES for SARS-CoV-2 became apparent. WES provided first evidence of importation of the virus into new geographical regions[17], heralded the onset of new waves of infection, illustrated disease transmission patterns and allowed inference of relative population burden during endemic phases[18]. WES also provided material for genotyping that demonstrated the presence of a broader range of variants than seen clinically[19]. In light of this global experience, the WHO issued updated guidance for countries conducting WES (WHO guidelines), citing the ability of WES to overcome the limitations of laboratory-based surveillance. However, despite these advantages and recommendations, there is a paucity of evidence integrating socioeconomic factors and disease epidemiology based on clinical testing data with wastewater surveillance in order to substantiate the claims that WES overcomes clinical testing limitations. Thus, definitive evidence for the ability of WES to overcome the limitations of laboratory-based surveillance is urgently required. We used clinical, wastewater surveillance, demographic and socioeconomic data in two different socio-economic contexts to explain and quantify the relationship between these variables and burden of SARS-CoV-2 disease, thus substantiating the use of WES as a necessary surveillance tool that enables interpretation of clinical surveillance data.

## Methods

### Conceptual framework

We developed a conceptual framework (Fig. 1) to demonstrate the relationships between wastewater concentrations of SARS-CoV-2 and SARS-CoV-2 disease indicators in sewered communities. The population burden of SARS-CoV-2 infections determines the levels of SARS-CoV-2 in wastewater. However, clinical indicators (including incidence rate, testing rate and proportion testing positive) reflect the distribution of testing, health care availability and accessibility. As these factors may be influenced by socio-economic conditions, reported case rates may not accurately reflect the true burden of infections. By determining the inter-relationships between wastewater surveillance data, social determinants of health and reported SARS-CoV-2 cases, the role of WES may be better understood.

**Fig. 1.**
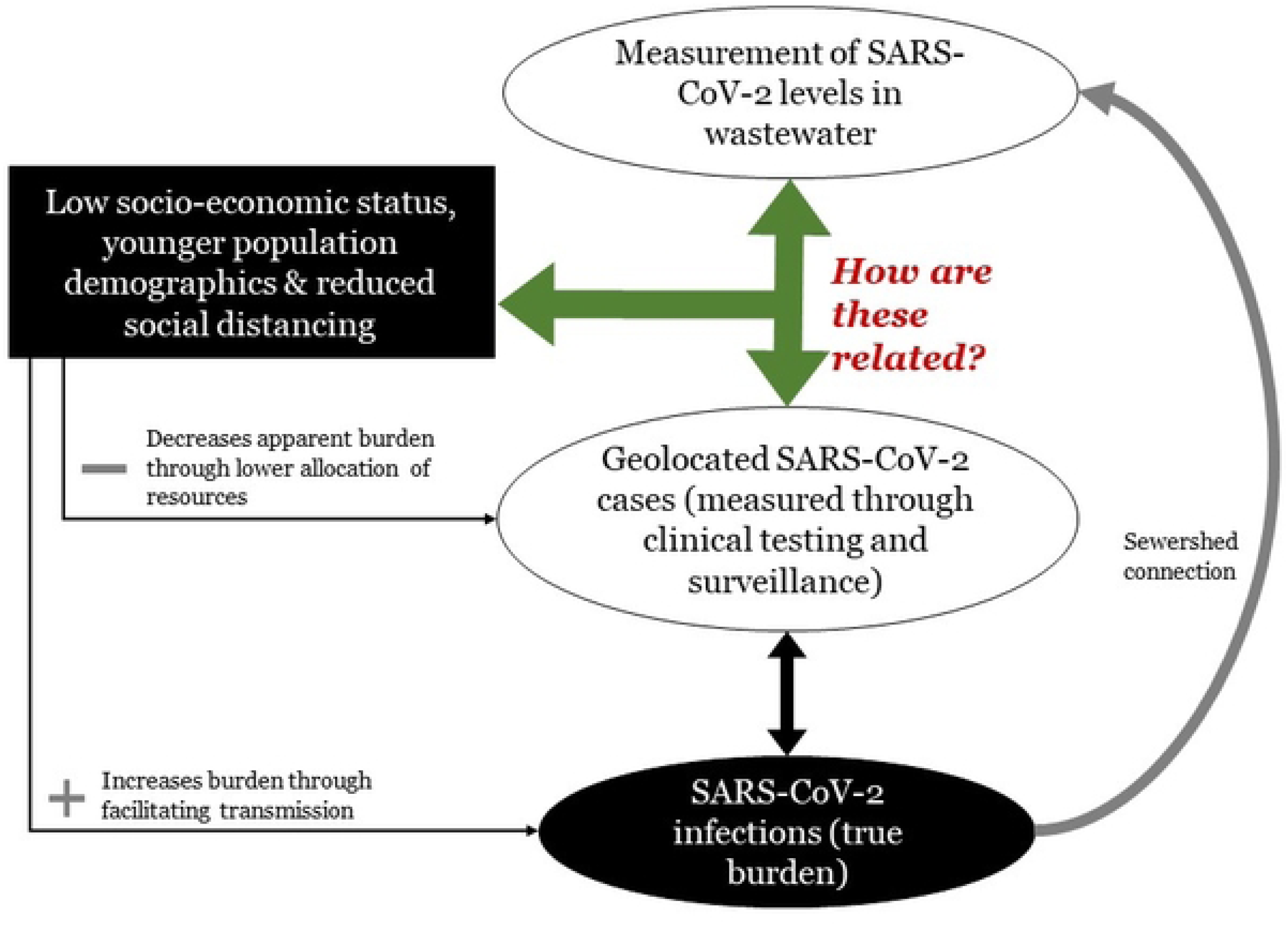
Diagram indicating the direction of influence of socio-economic status, population structure and mixing on the true burden of SARS-CoV-2 cases, reported burden of SARS-CoV-2 cases and levels of SARS-CoV-2 in wastewater. The green arrow indicates the research question posed by this work

### Study setting

The study took place in two sewersheds (sewershed D and sewershed O) in different metropolitan areas of Gauteng Province, South Africa. These sewersheds were purposively selected on the basis of geographical representativeness of city populations, namely sewershed D (City of Tshwane), to the north of the Gauteng Province, and sewershed O (City of Ekurhuleni) in the east of Gauteng Province (Fig. 2). Sewershed D is residential with formal housing in the west and central areas, and the metropolitan central business district to the east. A small area of informal housing exists to the far west. Sewershed O is mostly residential with low density, low-rise housing with areas of industrial and manufacturing activity to the north east. Informal settlements and backyard shacks are present in most neighbourhoods in the central areas, whilst a wealthier community of gated estates with a low population density is present in the northernmost section.

**Fig. 2.**
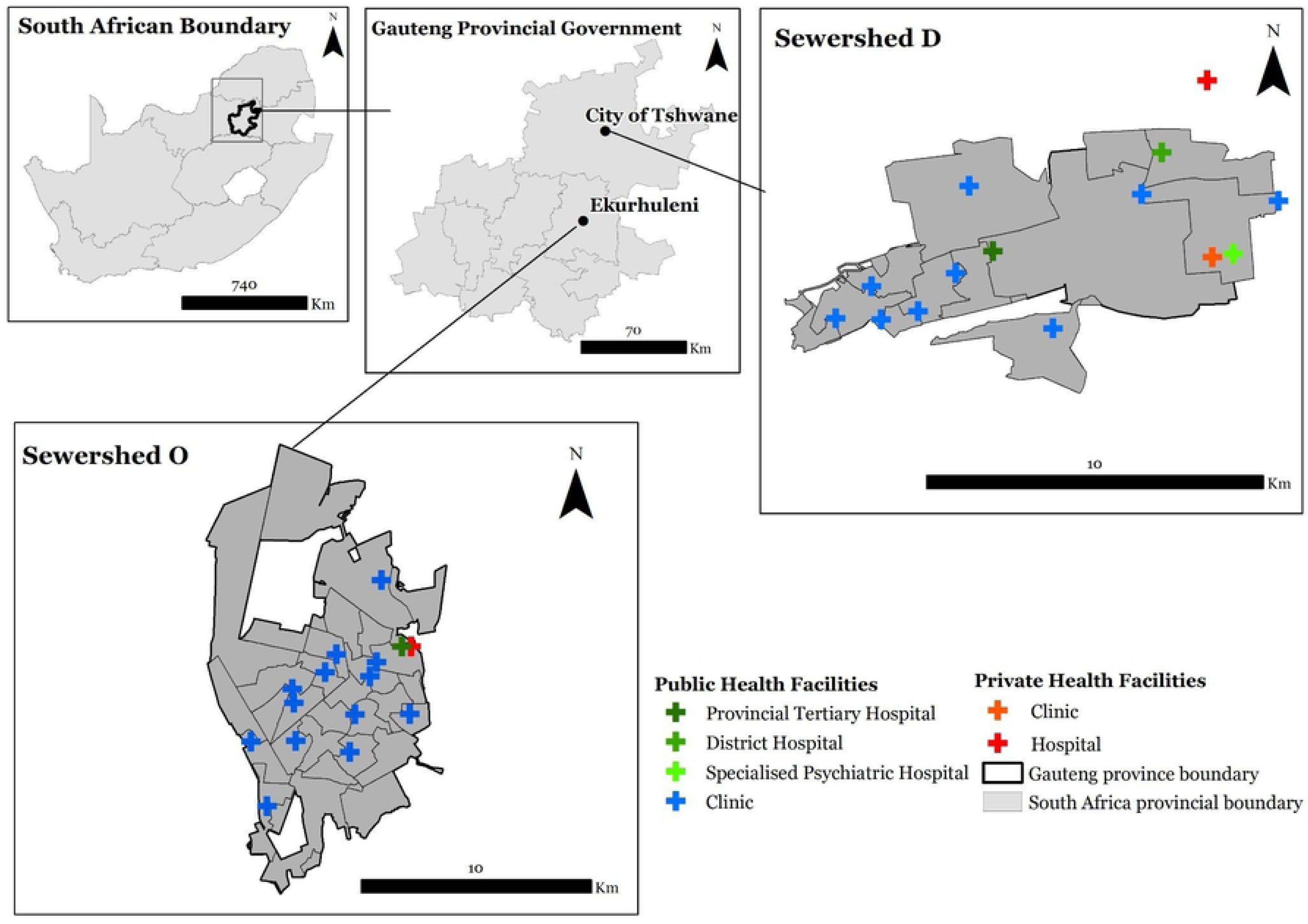
Map showing the spatial location of the two sewersheds and locations of health care facilities within the Gauteng Province

### Data sources and study period

#### Clinical SARS-CoV-2 laboratory testing and data management

In South Africa public and private laboratories were legally mandated to report all SARS-CoV-2 test results and patient data including residential address via the National Institute for Communicable Diseases (NICD). These data had been geocoded by NICD as part of outbreak response activities and all cases geocoded to Gauteng were extracted. Following ethics review and written approval for this study, anonymised and deidentified geocoded data for positive and negative SARS-CoV-2 PCR test results were supplied for the study period (1 June 2021 (epidemiological week 22 of 2021) to 18 March 2022 (epidemiological week 11 of 2022)) and the data was obtained on 23 May 2023.

#### Wastewater SARS-CoV-2 laboratory testing and data management

Routine SARS-CoV-2 wastewater surveillance data (obtained from laboratory processing of one litre grab-samples collected weekly as previously described[9]) from wastewater treatment plant (WWTP) ‘D’ and ‘O’ were identified for the study period. We obtained wastewater flow rates (in ML per day) from WWTP managers.

#### Quality of life survey-data

The Quality of Life (QoL) survey is a biennial household survey produced by the Gauteng City-Region Observatory (GCRO) which covers a range of topics including demographics, access to services and perceptions of residents. The survey is weighted to ensure representativity to ward level, a geopolitical division of municipalities, developed by the Municipal Demarcation Board and the smallest unit for which demographic and socioeconomic data are provided in South Africa. We obtained data from the 2020/21 (QoL6) survey (n=13,616 respondents) pertaining to 207 and 781 respondents from sewersheds D and O respectively[20].

#### Maps of sewershed reticulation methods

Data for the sewage networks and pipelines were obtained from the City of Tshwane and the City of Ekurhuleni, respectively. The City of Tshwane provided a comprehensive dataset including sewershed areas, manholes, and distribution pipelines. The City of Ekurhuleni consulting engineers provided shapefiles of the sewershed. ArcGIS 10.6.2 software was used for all mapping and spatial analysis.

#### GTI hexagon data for population demographics

Population demographic variables were extracted from a 2020/2021 dataset compiled by GeoTerraImage (GTI) comprising population estimates grouped by age cohorts in 400m sided hexagon (0.103755 km^2^).

### Data synthesis and analysis

#### General approach

We overlaid sewershed shape files, hexagonal population data, geolocated SARS-CoV-2 cases and geolocated QoL respondents in order to create a dataset comprising these elements for sewersheds D and O. To minimise the effect of the Modifiable Area Unit Problem (MAUP) that is encountered when polygon values require changing because of a change in the shape (zoning effect) or overall area (size effect) of the polygon, we scrutinised our datasets to determine the most viable polygon layer for all spatial datasets to allow for aggregation and comparability without disaggregation of polygon data. Ultimately we manipulated all data to the ward level allowing for comparative analysis over space and time. We conducted these analyses using SPSS software version 29.0.2.0 (20).

#### Data extraction and aggregation to determine population size, socio-economic and epidemiological parameters by sewershed

To determine population size and age structure within wards and sewersheds, we extracted population data from the GeoTerraImage (GTI) dataset for hexagons whose centroids fell within the sewershed and ward boundaries, and aggregated population data into four broad classes, namely children (0-4 years), adolescents (5-19), adults (20-59) and elderly (≥60 years). We geolocated QoL respondents within ward and sewershed boundaries and extracted and aggregated relevant socioeconomic and health fields for these individuals. Geolocated positive and negative SARS-CoV-2 PCR test data were aggregated by ward and sewershed level, and used together with population denominator data for each spatial unit to determine overall and weekly incidence, positivity rate and testing rate per 100,000. We created epidemiologic curves using Microsoft Excel 365 using incidence, positivity and testing rates together with SARS-CoV-2 concentration in wastewater for each catchment[21].

#### Principal Component Analysis (PCA)

We conducted principal component analysis (PCA) on the socioeconomic, demographic and clinical variables for each sewershed, using ward-level aggregated data as the unit of analysis to ascertain which variables relate most to epidemiological indicators in each sewershed. Using R (RStudio v4.0.2) and for each sewershed, we generated 1) a correlation coefficient (r) matrix for inter-variable correlation analysis; 2) a Scree plot for ascertaining the dimensions contributing the most to the explained variance as a percentage; 3) a variable loading graph for ascertaining the variables which contribute to the identity of the prominent dimensions; 4) and a biplot for plotting the variables as vectors in 2D space against the two most prominent dimensions.

#### Determination of theoretical infectious case load using mass balance equations and comparison with laboratory-confirmed cases by epidemiological week

We estimated the theoretical number of persons infected in each sewershed by epidemiological week using mass balance equations from Acheampeong *et al*[22], and Hoffman *et al*[23] as follows:

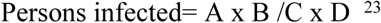

where A= RNA per L of wastewater (natural scale); B= estimated flow (L/day) obtained from the wastewater treatment authorities; C= grams of faeces/ person-day, estimated at 128g[24]; D= SARS-CoV-2 RNA per gram faeces, estimated at 2·58 x10^8 gene copies shed per day per infected person[23]. We assumed a linear regression to determine the number of persons infected per 100 laboratory-confirmed cases by epidemiological week within each sewershed.

### Ethical statement

This study was reviewed and approved by the University of the Witwatersrand Human Research Ethics Committee (HREC), M220904. In addition, the National Institute for Communicable Diseases obtained Ethics Approval for essential communicable disease surveillance and outbreak and response activities including SARS-CoV-2 (M210752).

## Results

### Demographic, socioeconomic and health characteristics

Sewershed O (land area =120 km^2^, population=905,996 persons) has around four times the number of people and just under double the population density of sewershed D (Table 1). Both sewersheds are dominated by younger, working age cohorts, with lower proportions of persons over 60 years of age. The proportion of households earning <USD90, together with the proportion relying on public transport, suggest that households in sewershed O are poorer. As many as 13% of households in sewershed O share sanitation, compared with around 2% in sewershed D. Regarding health care, responses suggested that up to 10% of households in sewershed O vs 4% in sewershed D struggled to access health care during 2020-2021 period. Up to 35% of households in Sewershed O were unable to maintain SARS-CoV-2 non-pharmaceutical measures.

**Table 1:** Comparative table of key SARS-COV-2, wastewater surveillance, socioeconomic and demographic variables for sewersheds D and O during the study period 1 June 2021 (epidemiological week 22 of 2021) to 18 March 2022 (epidemiological week 11 of 2022). Variable definitions are included.

|  | Sewershed D | Sewershed O |
| --- | --- | --- |
| **Sewershed area (km<sup>2</sup>)** | 62·0 | 120·7 |
| **Total population** | 245,935 | 905,996 |
| **Population density (persons/km<sup>2</sup>)** | 3·97 | 7·51 |
| **Number of wards for analysis** | 11 | 26 |
| **Demographic data (number of people, %)** |  |  |
| **0-4 years** | 21,196 (8·6) | 82,230 (9·1) |
| **5-19 years** | 55,965 (22·8) | 204,225 (22·5) |
| **20-59 years** | 150,798 (61·3) | 563,428 (62·2) |
| **≥60 years** | 17,976 (7·3) | 56,111 (6·2) |
| **Wastewater surveillance** |  |  |
| **Mean daily flow rate of wastewater through the treatment plant (ML, standard deviation)** | 38·4 (9·7) | 106·2 (9·9) |
| **Median (interquartile range) of SARS-CoV-2 (genome copies/mL)** | 47·2 (100·5) | 46·9 (109·7) |
| **SARS-CoV-2 laboratory surveillance data and indicators** |  |  |
| **Testing rate (tests/100 000 population)** | 23·7 | 10·6 |
| **Total recorded SARS-COV-2 cases during study period** | 11,026 | 15,293 |
| **Mean positivity rate (% , sd)** | 17 (12·8) | 14 (9·9) |
| **Cumulative incidence rate (cases /100 000 population)** | 4,483·3 | 1,688·0 |
| **Socio-economic and health indicators from Quality of Life Survey 6 (2020/2021)** |  |  |
| **Income below R1600 (sd)**<br>*Percentage of households that have a combined income less than R1600 per month* | 21%(4·6) | 33%(4·6) |
| **Reliance on public transport (sd)**<br>*Percentage of respondents that rely on public transport* | 40%(0·5) | 47%(0·5) |
| **Access to shared sanitation (sd)**<br>*Percentage of respondents who generally use communal toilets or toilets not connected to the sewage system (e.g. portable toilets)* | 2%(0·2) | 13%(0·3) |
| **Quality of Life score (sd)**<br>*Score out of 100. Calculated with 33 variables, grouped in 7 dimensions* | 63(9·0) | 59(11·0) |
| **Struggled access to health care (sd)**<br>*Percentage of respondents that struggled to access healthcare from March 2020 to October 2021* | 4%(0·2) | 10%(0·3) |
| **Refused COVID-19 testing (sd)**<br>*Proportion of respondents who tried but were denied access a COVID-19 test between March 2020 and October 2021* | 5%(0·2) | 3%(0·2) |
| **COVID-19 Index (sd)**<br>*Percentage respondents unable to implement SARS-CoV-2 preventative measures including social distancing* | 29(19·0) | 35(20·0) |

(sd)= standard deviation, km=kilometre

### SARS-CoV-2 clinical testing, incidence rates and percentage test positive (PTP)

During the study period (1 June 2021 to 18 March 2022), 78% and 60% of positive and negative test results respectively were successfully geolocated. The SARS-CoV-2 clinical testing rate per 100,000 in sewershed D (23.7) was over twice the rate in sewershed O (10.6). Despite higher absolute numbers of SARS-CoV-2 cases, sewershed O had a cumulative incidence of laboratory-confirmed SARS-CoV-2 cases around three times lower than sewershed D (15,293 total cases and 1,688/100,000, vs 11,026 total cases and 4,483/100,000) (Table 1 and Fig. 3).

**Fig. 3.**
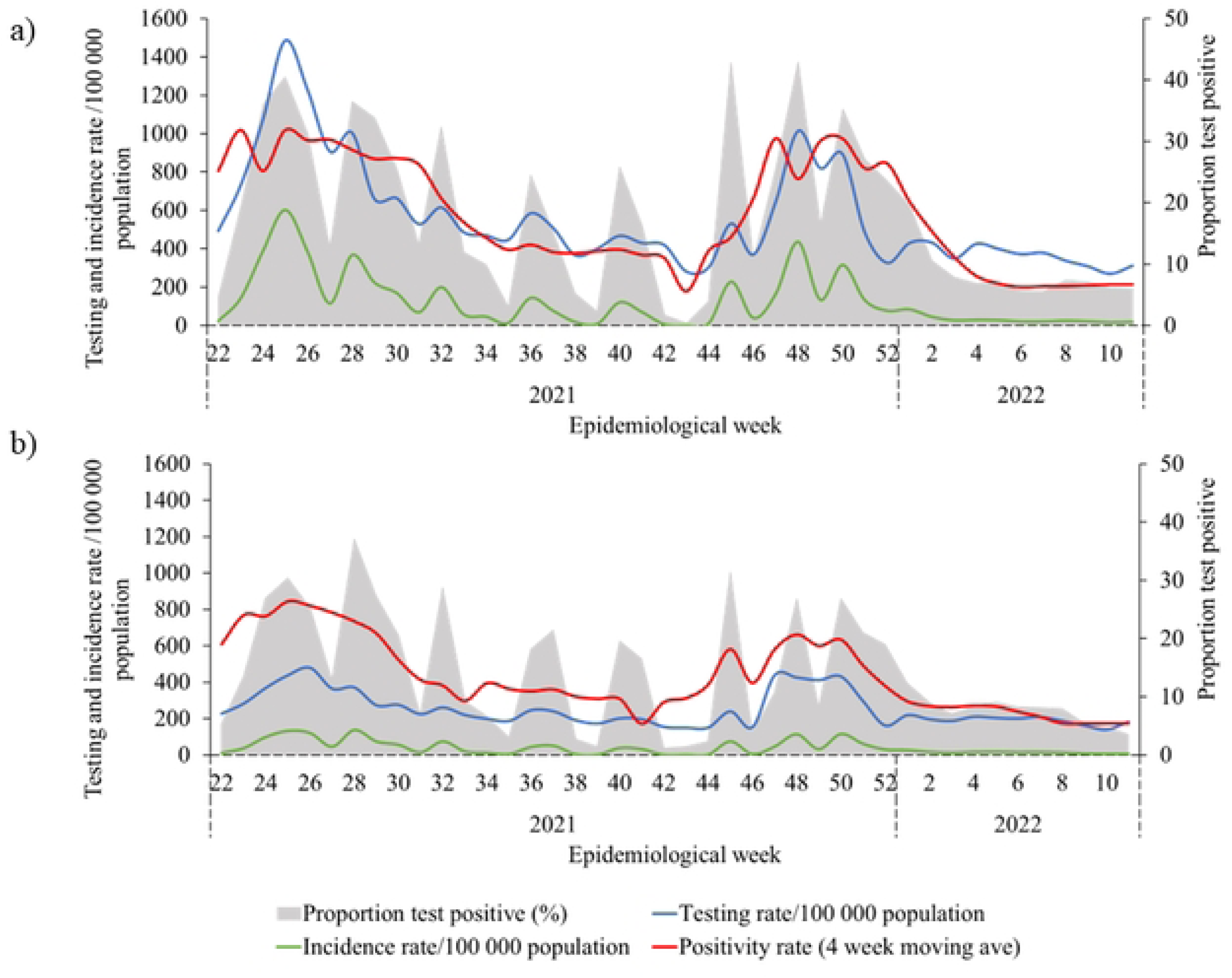
SARS-CoV-2 testing rate (per 100 000 population), incidence rate (per 100 000 population), and 4 week moving average proportion test positive (%) by epidemiological week 22, 2021 to week 10, 2022, for (a) sewershed D and (b) sewershed O.

Fig. 3 illustrates changes in testing, incidence and proportion testing positive (PTP) by epidemiological week during the two waves of infection that occurred during the study period, namely Delta (between epidemiological weeks 15 and 40 in 2021) and Omicron (epidemiological weeks 46 - 51 in 2021).Whilst all indicators follow the similar trends, rates are lower in sewershed O.

### Relationship between clinical case data and socioeconomic status

Correlation matrices for sewersheds D and O reveal that clinical indicators (PTP, cumulative incidence and testing rates) do not exhibit consistent relationships with demographic, or socio-economic indicators within and between sewersheds (Fig. 4). In sewershed O, *PTP* correlates with indicators associated with poverty (*access to* and *use of shared sanitation*, *% of the population with income below USD 90*, and *COVID-19 index* (% unable to implement COVID-19 preventive measures)), whilst this relationship is not evident in the more affluent sewershed D. In sewershed D, it appears that PTP and cumulative incidence are correlated with *quality of life, use of public transport* and *proportion of persons with income below USD 90*, suggesting that economic conditions influence access to testing, in turn leading to apparently low cumulative incidence. Taken together, in areas with poorer socio-economic conditions, testing rates and cumulative incidence may not reflect disease burden, whilst PTP better reflects disease risk.

**Fig. 4.**
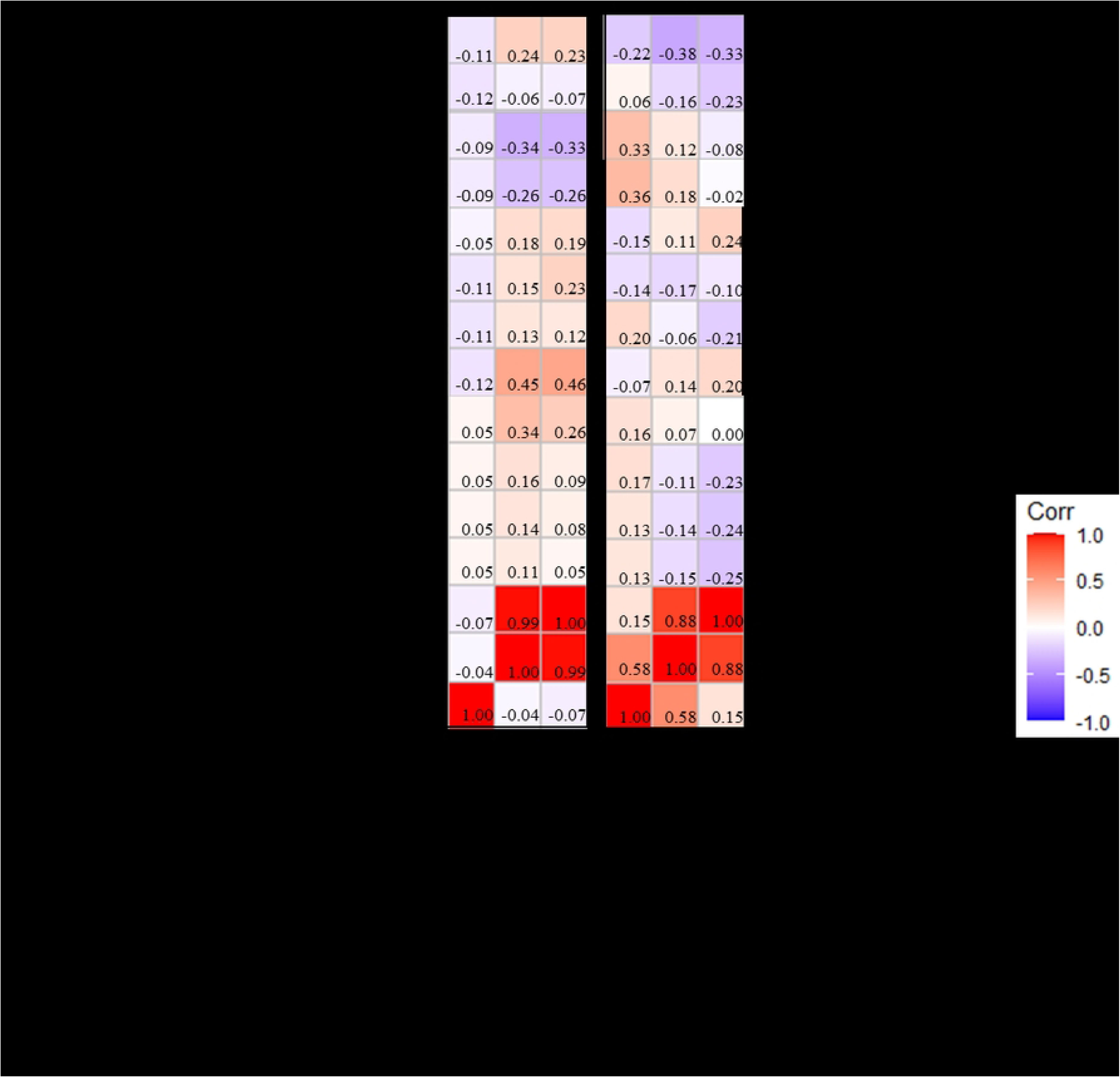
Truncated correlation matrices between socio-economic and demographic parameters against testing, cumulative incidence and mean proportion test positive for sewersheds D and O, annotated as a ‘heat map’ to represents the Spearman’s correlation coefficient (r) between socio-economic, demographic variables vs clinical variables (1 June 2021 to 18 March 2022).

Scree plots for sewershed D and O demonstrate that 76.7% and 77.5% of data are explained by dimensions 1 and 2 (Fig. S1). The variable loading plot for sewershed D indicates that *access to shared sanitation*, *reliance on non-sewered toilets* and p*opulation groups under 60 years of age* contributed the most to composition of dimensions 1 and 2 (Fig. S2a) whereas in sewershed O, population group *under 60 years of age*, *testing rate* and *cumulative incidence rate* contributed the most to the composition of dimension 1 and 2 (Fig. S2b). In both sewersheds, testing rate and cumulative incidence contributed to dimensions 1 and 2, but the *proportion test positive* and *refused COVID testing* variables contributed minimally.

The biplots (Fig. 5) demonstrate that SARS-COV-2 testing and incidence rates contribute equivalent influence and spatial distribution on socioeconomic factors for both sewersheds, whilst the *proportion test positive* indicator was much less affected by socioeconomic factors. In the biplot for sewershed D, *access to healthcare* and *refused access to COVID testing* are more associated with cumulative incidence and testing rates, while only *refused access to COVID test* is associated with the two clinical variables in sewershed O. In sewershed D, the clustering of the *Quality of Life score* and the *greater than 60 years* age group as well as the clustering of *COVID-19 Index* with *income below USD90* indicated these variables are influenced by and can inform one another. In sewershed O, *struggled to access health care* and *Quality of Life score* illustrated similar clustering with the *COVID-19 Index* and *access to public transport*.

**Fig. 5.**
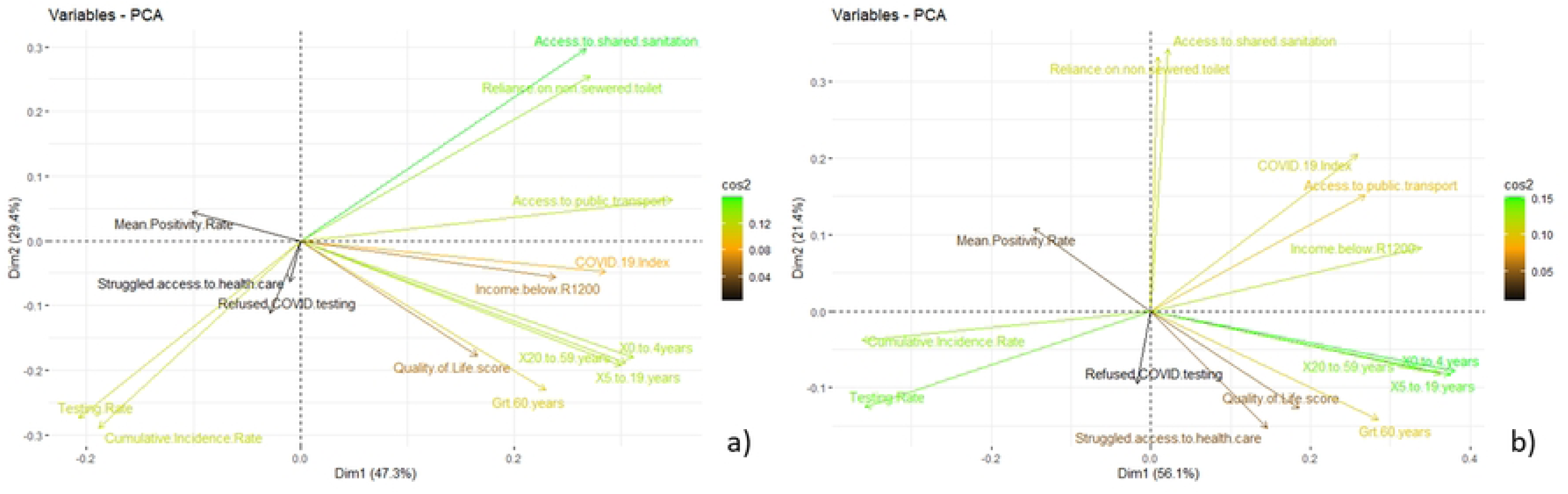
PCA biplots displaying socioeconomic and demographic status parameters, cumulative incidence rate, testing rate and mean positivity rate within (a) sewershed D and (b) sewershed O. In the biplots, the magnitude and colouring of the vectors are related to the variable loading scores, while the vector direction and quadrant location is informed by the interrelationship between variables and their contribution to dimensions 1 and 2.

### Quantitative SARS-CoV-2 surveillance in wastewater

Wastewater concentrations of SARS-CoV-2 and geolocated laboratory-confirmed SARS-CoV-2 cases in sewersheds D and O were significantly correlated in each sewershed, though less so in sewershed O (Spearman’s correlation coefficient 0·723 (p<0.001)), 0.476 (p=0·020) for sewersheds D and O respectively). In each sewershed, higher concentrations and case-loads were observable during the Delta and Omicron waves that occurred during the study period (Fig. 6) and both reached an ebb during weeks 40-42 of 2021. Wastewater concentrations (measured in log genome copies per millilitre) for both sewersheds ranged between 0·5 and 3·5 log copies/mL and were at similar concentrations in the same epidemiological week. Weekly laboratory-confirmed case-counts followed similar trends despite different population sizes in the two catchments.

**Fig. 6.**
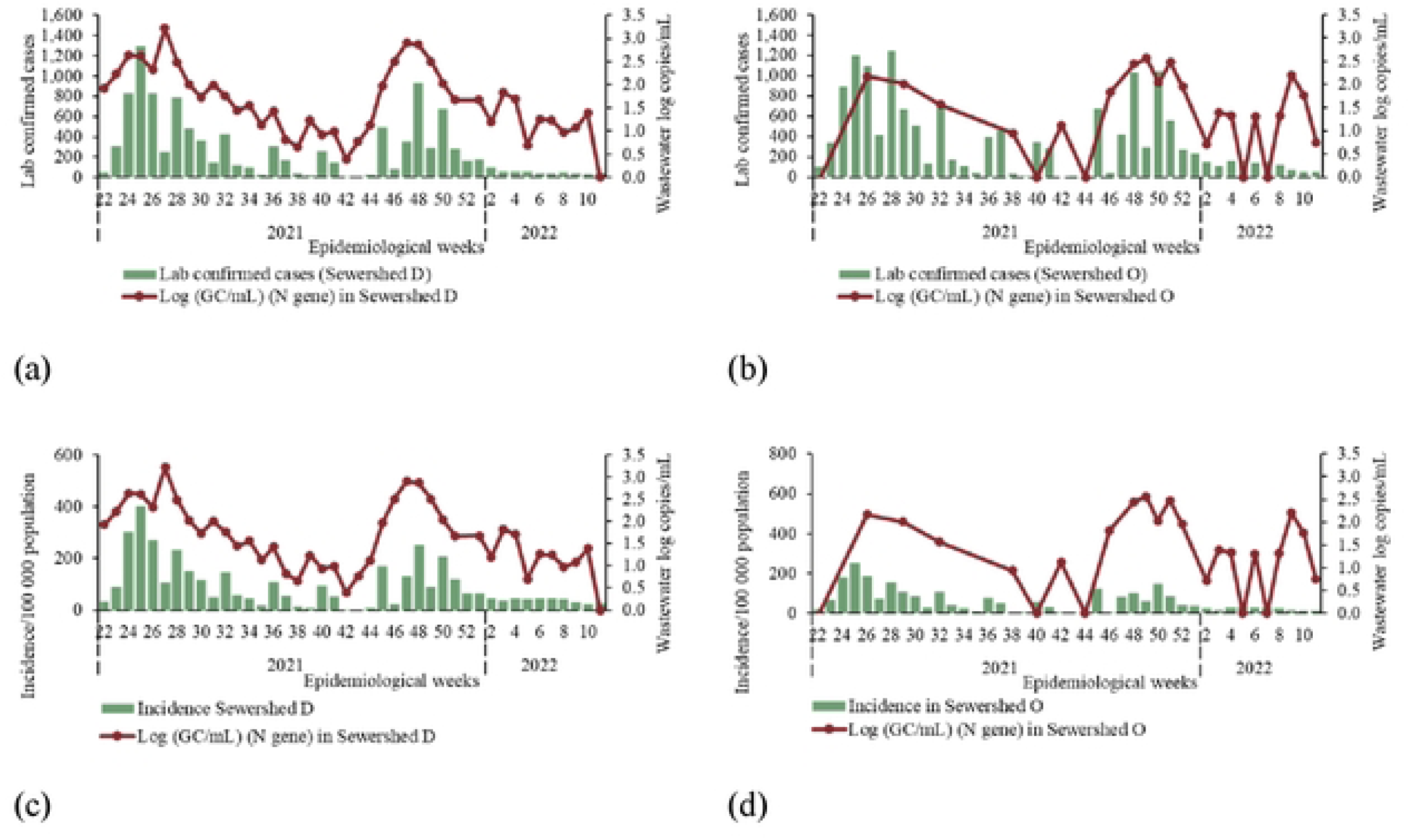
SARS-CoV-2 concentrations in wastewater in log-transformed genome copies per millilitre (right axis) and the number of laboratory-confirmed cases (top figures, green bars) or incidence per 100,000 persons (bottom figures, blue bars) of SARS-CoV-2 geolocated to a residential address in the sewershed by epidemiological week from week 22, 2021 to week 10, 2022 for sewersheds D (left, figures a and c respectively) and O (right, figures b and d respectively).

### Comparison of estimated and actual SARS-CoV-2 case burden

Regression analysis by sewershed of laboratory-confirmed SARS-CoV-2 cases versus theoretical number of infections indicated that for each 100 reported cases, sewershed O likely had over 63,000 infections compared with sewershed O, with 2,700 cases (Fig. 7).

**Fig. 7.**
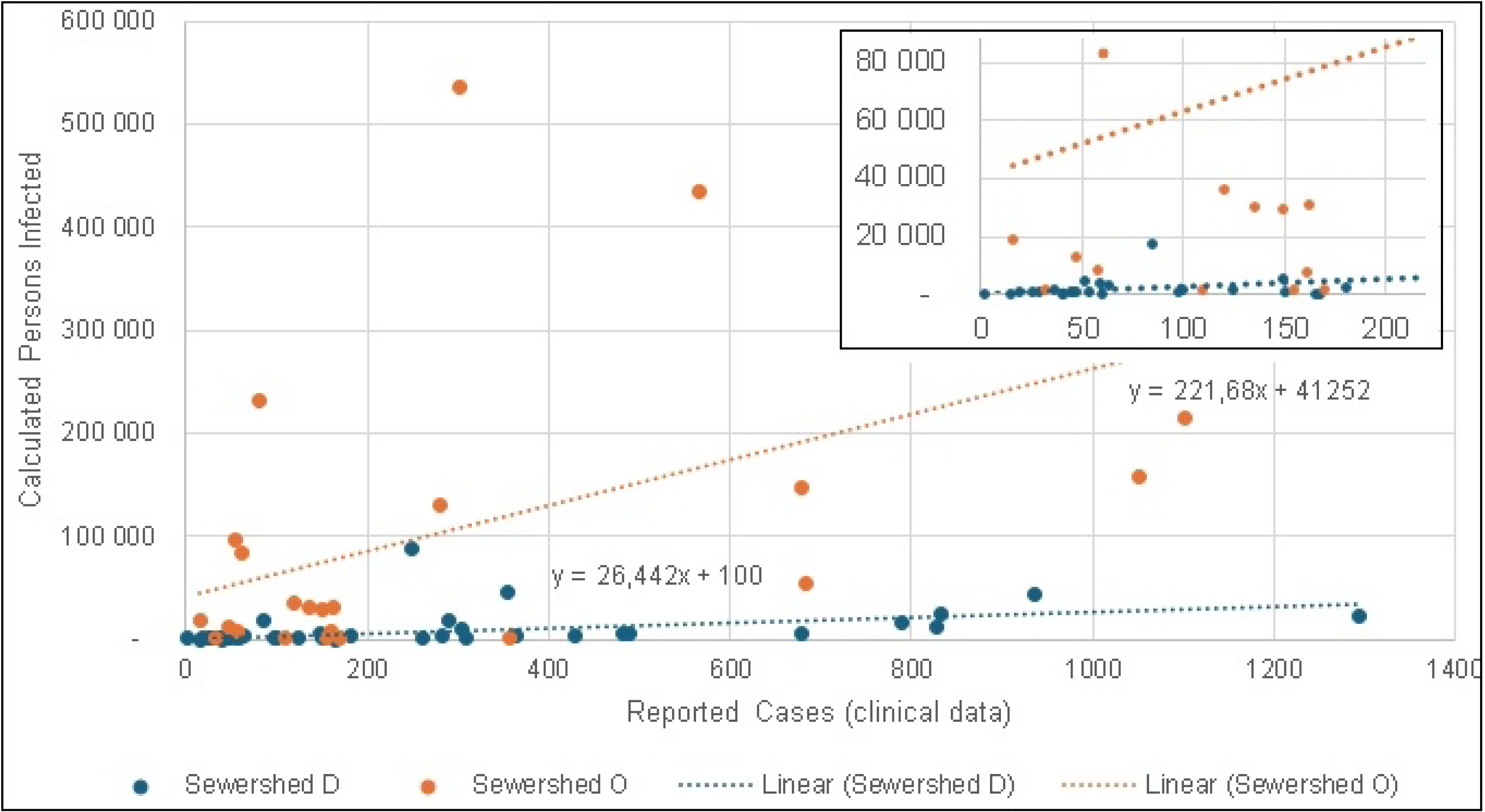
SARS-CoV-2 concentrations in wastewater in log-transformed genome copies per millilitre (right axis) and the number of laboratory-confirmed cases (top figures, green bars) or incidence per 100,000 persons (bottom figures, blue bars) of SARS-CoV-2 geolocated to a residential address in the sewershed by epidemiological week from week 22, 2021 to week 10, 2022 for sewersheds D (left, figures a and c respectively) and O (right, figures b and d respectively).

## Discussion

In our transdisciplinary spatial analysis of clinical and environmental data during two large COVID-19 pandemic waves in sewersheds with differing socioeconomic conditions, we observed that despite different population sizes, the concentrations of SARS-CoV-2 in wastewater and the absolute numbers of SARS-CoV-2 cases by epidemiological week were similar. In the light of assumed equivalent excretion rates of SARS-CoV-2 in infected individuals and identical demographic profiles in each sewershed, equivalent wastewater concentrations suggest vast under-reporting of cases in the poorer sewershed. Our socioeconomic analysis demonstrated negative correlations between income and SARS-CoV-2 cumulative incidence and testing rates in the poorer sewershed. Overall, we demonstrated the co-contributory influences of socio-economic indicators on access to SARS-CoV-2 testing and cumulative incidence, thus reflecting that apparent incidence rates mirror access to testing and socioeconomic considerations rather than true disease epidemiology. These analyses demonstrate how WES provides valuable information to contextualise and interpret laboratory-based epidemiological data.

Laboratory-based surveillance systems under-represent the true burden of disease due to a combination of asymptomatic infection, individual and cultural practices regarding health seeking, quality of health care and socio-economic factors that impair access to testing. Furthermore, these same socio-economic factors including education and poverty, are associated with higher SARS-CoV-2 rates[6]. Underreporting of SARS-CoV-2 cases in South Africa was evident through excess mortality reports which indicated over 70,000 excess deaths vs 28,000 reported COVID-19 deaths during 2020[13]. Households in lower income groups, those who rely on public health care, and Black African and Coloured population groups were more likely to have struggled to access healthcare and testing facilities[25]. Our data, demonstrating that cumulative incidence and testing rate in sewershed O was negatively correlated with low income (< USD 90 household income per month) suggest that clinical testing was missing this population segment.

Few studies have triangulated clinical testing, wastewater surveillance data and socio-economic factors. Using WES data in contrast to clinical testing data, Lancaster *et al*[26] identified specific communities (Black, poor) that were more vulnerable to SARS-CoV-2. Saingam *et al*[27] demonstrated that a machine learning model to predict COVID-19 and post-infectious sequelae is strengthened when including WES data together with socio-demographic parameters. Rogawski McQuade *et al*[28] in Bangladesh, using purposively selected sampling sites across the economic spectrum, demonstrated equivalent SARS-CoV-2 levels in wastewater, but 23 and 70 times the number of clinical tests conducted in the wealthier vs middle and poor income areas. Our interdisciplinary study is the first to use WES data to show evidence for underreporting of SARS-CoV-2 in the context of measurable and statistically significant associations between economic conditions and SARS-CoV-2 incidence and testing rates, something which has been intuitively speculated but not empirically demonstrated[29].

Several limitations exist in our data collection and interpretation. A small proportion of SARS-CoV-2 cases were not successfully geolocated, however non-geolocated cases are more likely to have originated outside the province, or from persons resident in informal settings thus not contributing to wastewater levels of SARS-CoV-2. The absence of wastewater sampling points within wards precluded inclusion of SARS-CoV-2 levels in PCA analyses. Whilst our wastewater data was generated from urban sewersheds, similar findings in non-sewered areas suggest that our findings are generalisable across sewered and non-sewered settings[28].

The addition of WES data to national and global surveillance systems will strengthen sensitivity of event detection for outbreak and pandemic disease, monitoring of endemic disease trends, and will jointly provide material for genomic epidemiology[19]. An evaluation of detection of H5N1 avian influenza by six northern hemisphere surveillance systems in 2010 demonstrated an increase in sensitivity of detection using a combination of data inputs. Authors concluded that the range of surveillance methodologies and variation in system designs created synergy between systems, led to improved data quality and validity, and allowed data to converge on event detection[30]. In a low-middle income country, the need for multiple surveillance systems is even more necessary, as data quality and completeness from single modality surveillance systems may vary, leading to challenges in decision making during a crisis.

Our findings provide evidence to support intuitive thinking that WES overcomes testing biases particularly in situations with socio-economic disparities and weaker clinical disease surveillance programmes for SARS-CoV-2. Our findings are likely broadly applicable to all communicable disease surveillance programmes, as biases affecting clinical surveillance programmes are not disease-specific, and all pathogens infecting humans are likely to find their way into wastewater albeit in varying quantities. As such, our findings strengthen the case for investment in implementation of WES. Ongoing implementation of WES will allow public health authorities to determine optimal configurations of WES surveillance systems for each pathogen and public health use-case. Further research to determine optimal sample collection, processing and testing methods is needed.

Interpretive frameworks or mathematical models will support integration and interpretation of WES data with clinical surveillance data.

## Data Availability

All data produced in the present study are available upon reasonable request to the authors. Data from the GCRO Quality of Life survey is freely available under the CC BY 4.0 licence from the DataFirst service and available online at https://www.datafirst.uct.ac.za/dataportal/index.php/catalog/GCRO/about

https://wastewater.nicd.ac.za/

## Data sharing

Clinical and laboratory data were generated at the National Institute for Communicable Diseases. The Quality of Life survey dataset is freely available under the CC BY 4.0 licence from the DataFirst service. For more information please. Derived data supporting the findings of this study are available from the corresponding author GM on request.

## Acknowledgements

The authors would like to thank the following groups – namely the NICD IT and Surveillance Data Warehouse staff for provision of geolocated SARS-CoV-2 case data; the NICD Epidemiology and Surveillance teams that curated SARS-CoV-2 databases during the pandemic; and the City of Tshwane and City of Ekurhuleni water and sanitation departments for provision of sewershed maps.

## Author contributions

GM, FE, YN, LN, SR, MY and KM were involved in the conception design of the study. YN, LN, GM and FE curated the data. GM, FE, YN, PM, KO and LN completed the formal analysis. PM and FE completed data visualisation. GM wrote the original draft. MM, NN, SR and MY were involved in laboratory testing. SM did the project administration. MY and KM obtained funding for this study.

All authors critically reviewed the manuscript and approved the final version.

