## Supplementary material for "Wastewater surveillance overcomes socio-economic limitations of laboratory-based surveillance when monitoring disease transmission: the South African experience during the COVID-19 pandemic"

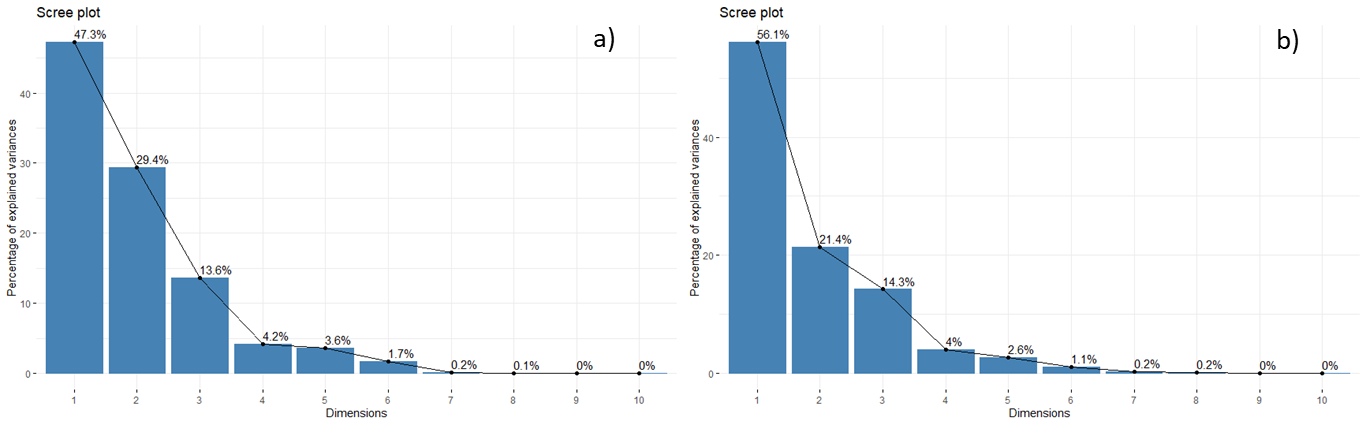

**Fig. S1. Principal Component Analysis Scree plots indicating the percentage of variance explained by principal components. Dimension 1 and 2 account for 76.7% in sewershed D (a) and 77.5% sewershed O (b).**

In both sewersheds, dimensions 1 and 2 explained three quarters of the variance within the respective datasets (77% of the variation in sewershed D and 78% of the variation in sewershed O). Dimensions 4 to 10, in both sewersheds, explained less than 10% of the total variance in both datasets. The variable loadings plot and the biplots were thus created for the comparison of dimensions 1 and 2 for both sewersheds.

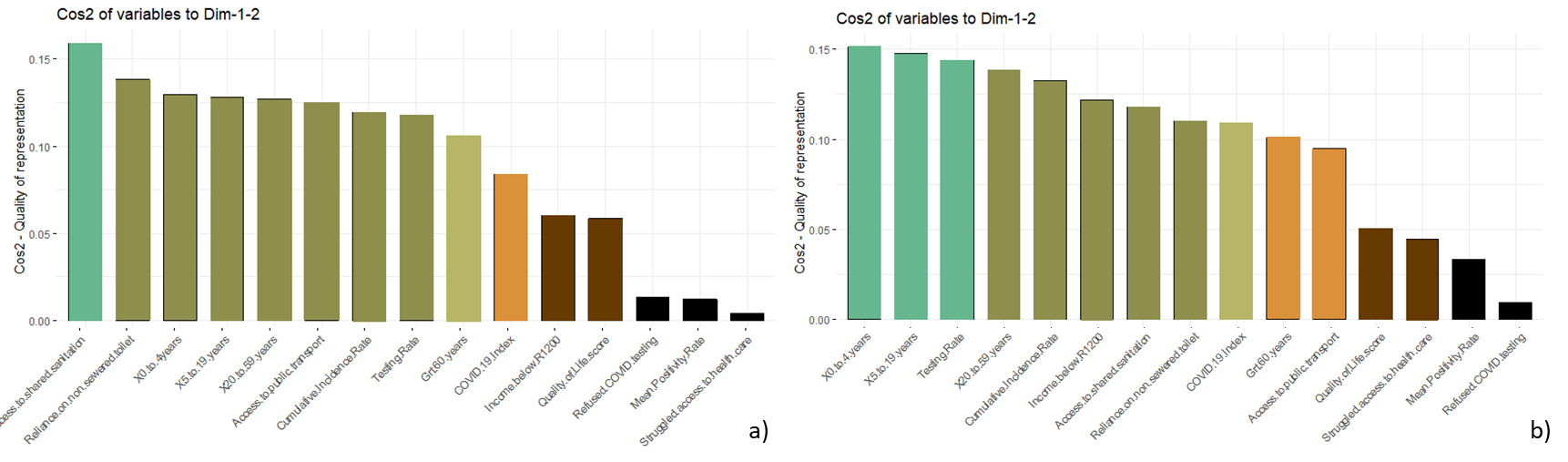

**Fig. S2. Variable loading bar plots of the variables contributing to the composition of dimensions 1 and 2 for (a) sewershed D and (b) sewershed O. Note: Colour ramp matches the PCA plots and the darker the shade the lower the loading value**
